# Supportive Care for Corona virus Pneumonia and Risk of Infection to Health Care Workers; A Rapid GRADE Review

**DOI:** 10.1101/2020.07.06.20146712

**Authors:** TK Luqman-Arafath, Sandeep S. Jubbal, Elakkat D Gireesh, Jyothi Margapuri, Hanumantha Rao Jogu, Hitesh Patni, Tyler Thompson, Suma Menon, Arsh Patel, Amirahwaty Abdulla, Sudheer Penupolu

**Affiliations:** Department of Internal Medicine, Wake Forest School of Medicine, Winston Salem, NC, USA; Department of Internal Medicine, University of Massachusetts Medical School, Worcester, MA, USA; Department of Neurology, Advent Health, Orlando, FL, USA; Department of Pulmonary Critical Care Medicine, Geisinger Medical Center, Danville, PA, USA; Florida Nephrology PLC, Orlando, FL, USA

## Abstract

**Background:** Avenues of treatment currently implemented for Covid-19 pandemic are largely supportive in nature. Non-availability of an effective antiviral treatment makes supportive care for acute hypoxic respiratory failure is the most crucial intervention. Highly contagious nature of Covid-19 had created stress and confusion among front line Health Care Workers (HCWs) regarding infectious risk of supportive interventions and best preventive strategies.

**Purpose:** To analyze and summarize key evidence from published literature exploring the risk of transmission of Covid-19 related to common supportive care interventions in hospitalized patients and effectiveness of currently used preventive measures in hospital setting.

**Data Sources:** Curated Covid-19 literature from NCBI Computational Biology Branch, Embase and Ovid till May 20,2020.Longitudinal and reference search till June 28,2020

**Study Selection:** Studies pertaining to risk of infection to HCWs providing standard supportive care of hospitalized Covid-19 mainly focusing on respiratory support interventions. Indirect studies from SARS,MERS or other ARDS pathology caused by infectious agents based on reference tracking and snow ball search. Clinical, Healthy volunteer and mechanistic studies were included. Two authors independently screened studies for traditional respiratory supportive-care (Hypoxia management, ventilatory support and pulmonary toileting) related transmission of viral or bacterial pneumonia to HCWs.

**Data Extraction:** Two authors (TK and SP) independently screened articles and verified for consensus. Quality of studies and level of evidence was assessed using Oxford Center for Evidence Based Medicine (OCEBM), Newcastle - Ottawa quality assessment Scale for observational studies and Grading of Recommendations Assessment, Development and Evaluation (GRADE) system for grading evidence.

**Data Synthesis:** 22 studies were eligible for inclusion. In 11 mechanistic studies, 7 were manikin based,1 was in the setting of GNB pneumonia, 2 were healthy volunteer study and 1 was heterogenous setting.Out of 11 clinical studies, 5 were case controlled and 6 were cohort studies. Risk of corona virus transmission was significantly high in HCWs performing or assisting endotracheal intubation or contact with respiratory secretion.(Moderate certainty evidence, GRADE B) Safety of nebulization treatment in corona virus pneumonia patients are questionable(Low certainty evidence, GRADE C).Very low certainty evidence exist for risk of transmission with conventional HFNC (GRADE D) and NIV (GRADE D),CPR (GRADE D),Bag and mask ventilation(GRADE D).Moderate certainty evidence exist for protective effect of wearing a multilayered mask, gown, eye protection and formal training for PPE use (GRADE B).Low certainty evidence exist for transmission risk with bag and mask ventilation, suctioning before and after intubation and prolonged exposure (GRADE C).Certainty of evidence for wearing gloves,post exposure hand washing and wearing N 95 mask is low(GRADE C).

**Limitations:** This study was limited to articles with English abstract. Highly dynamic nature of body of literature related to Covid-19, frequent updates were necessary even during preparation of manuscript and longitudinal search was continued even after finalizing initial search. Due to the heterogeneity and broad nature of the search protocol, quantitative comparisons regarding the effectiveness of included management strategies could not be performed. Direct evidence was limited due to poor quality and non-comparative nature of available Covid-19 reporting.

**Conclusions:** Major risk factors for transmission of corona virus infection were, performing or assisting endotracheal intubation and contact with respiratory secretion. Risk of transmission with HFNC or NIV can be significantly decreased by helmet interface, modified exhalation circuit or placing a properly fitting face mask over patient interface of HFNC. Evidence for risk of transmission with CPR, suctioning before or after intubation or bag and mask ventilation of very low certainty. Significant protective factors are Formal training for PPE use, consistently wearing mask, gown and eye protection.

## Introduction

After the 1918 Spanish flu, no other infectious disease has inflicted such a significant impact to the existence of humanity as the Coronavirus Disease of 2019 (COVID-19). It is caused by infection with Severe Acute Respiratory Syndrome Coronavirus – 2 (SARS CoV 2), which is a single stranded positive sense RNA virus belonging to the Coronaviridae family(1, 2). SARS-COV-2 is highly contagious compared to SARS or MERS virus and has a significantly higher mortality in comparison to common respiratory viruses like Influenza (2, 3). Clinical scientists, bench researchers, and exasperated front-line providers are racing against time to come up with effective treatment strategies for COVID-19. Supportive management of Acute Respiratory Distress Syndrome constitutes the mainstay of care of COVID-19. Majority of the patients infected with SARS-COV-2 (approx. 80%) develop only mild symptoms (5) and will likely not require medical attention. Priority in such cases should be for isolation to contain/mitigate virus transmission. But 14% will develop severe illness requiring oxygen therapy and approximately 5% will require intensive care unit treatment and often with mechanical ventilation support(4-6).Pulmonary complications are arguably the most common pathology noted in COVID-19.Guidelines vary considerably in their recommendations on supportive care and direct evidence is scarce. Multiple investigational approaches have been employed, particularly for the sickest of these patients and are primarily based on either extrapolation of data from SARS or MERS treatment strategies, in-vitro study results, or recently available clinical studies with relatively small sample sizes. Myriads of potential treatment strategies explored in COVID-19 patients are not supported by direct evidence, since data from clinical trials involving Covid-19 patients are still emerging. Major dilemma faced by Healthcare workers (HCWs) is risk of nosocomial transmission of Covid-19. SARS-CoV-2 is considered to primarily be spread by droplet transmission and direct contact even though there are some concern for transmission through aerosols(7).Studies based on treatment of ARDS caused by closely related corona viruses in the past (SARS or MERS) suggested commonly employed respiratory supportive care measures like intubation, Non-invasive ventilation (NIV),nebulization, Chest Physio Therapy and High Flow Nasal Cannula (HFNC) carry variable risk of transmission to HCWs(7). Past experience from SARS and MERS epidemics suggestive of almost 20-56 % infection rate in HCWs (8-11). During February 12–April 9, among 315,531 COVID-19 cases reported to CDC using a standardized form, 49,370 (16%) included data on whether the patient was a health care worker in the United States; including 9,282 (19%) who were identified as HCW (12).Incidence or risk factors of Covid-19 (symptomatic or asymptomatic) in HCWs are not available till date. In this review, we are appraising available evidence on risk of transmission of Covid-19 to HCW associated with commonly employed respiratory supportive care.

## Methods

Clinical questions were developed into a PICO format (Population, Intervention, Comparison, Outcomes). We attempted to adhere with Cochrane guidelines for systematic review when possible (13).Initial attempt was to gather evidence from direct studies related to Covid-19 infection,but initial screening of data bases(Medline,Embase and LitCovid) showed only 1 study involving Covid-19.Extensive reference search of multiple guidelines and expert commentaries was done to extent the search domain to include nosocomial transmission of respiratory infections including SARS and MERS.

(Two authors independently reviewed (SP and TK) a dynamic collection of articles related to COVID-19 from three databases namely, LitCovid literature collection, Embase and Ovid Medline. Comprehensive Search was conducted between May 2, 2020 and finalized on May 20, 2020. Longitudinal Search on LitCovid data base was continued till June 26, 2020 for inclusion of additional potentially relevant articles (1 study was included). LitCovid is a curated collection published on COVID-19 and updated on daily basis. This resource is provided free of charge to all researchers by NCBI Computational Biology Branch. LitCovid identified 35% more relevant articles than conventional keyword-based searches for entries such as ‘COVID-19’ or ‘nCOV’(14). A comprehensive search of Ovid Medline using a broad search terms pertaining to treatment aspect of Covid-19(See appendix 1 and Appendix 2 for details) was conducted with the general assistance of Librarian (Carpenter Library, Wake Forest University School of Medicine).This search was later transferred to Emabse data base. Ovid and Embase Search yielded 3751 and 1383 articles respectively. On the day this search was finalized on LitCovid data base(06/26/2020) there was a total of 25742 articles related to Covid-19 and upon further narrowing to subgroups ‘Treatment” and “Transmission”yielded a total of 6456 articles. Citations were managed with the help of EndNote X9 version software and duplicates were removed. A total of 4295 articles were retrieved including 75 articles from reference / snowball search. Articles were further organized into groups of interventions and assessed for inclusion by authors TK and SP. Articles not pertaining to respiratory supportive care(high flow nasal canula (HFNC), non-invasive positive pressure ventilation (NIV), proning, Chest physio therapy or other pulmonary toileting) were excluded. Following PRISMA like protocol other exclusions were Non-Adult, Animal studies,Editorial, Commentary, Letters without research data, systematic reviews, metanalysis, press releases, case reports, interviews, or conference proceedings. Articles pertaining to general supportive care were separated from specific pharmacologic treatment(part of another review being authored by same authors). Updates in literature were longitudinally monitored by authors JM and TJ. Following PRISMA (Preferred Reporting Items for Systematic reviews and Meta-Analyses) like protocol a total of 22 studies were included in the present study. Detailed overview of the systematic review process(15) displayed in **Figure 1**.Included studies with English abstracts were assessed for quality/level of evidence according to the guidelines and recommendations provided by the Oxford Center (Appendix.3)for Evidence Based Medicine and Newcastle - Ottawa quality assessment Scale(NOS-Appendix 4) for observational studies(16). Attempted to grade the evidence on harm or benefit of specific intervention when appropriate, following the GRADE (Grading of Recommendations Assessment, Development and Evaluation) approach(17). Individual study characteristics are given **in Table. 1** and **Table.2**

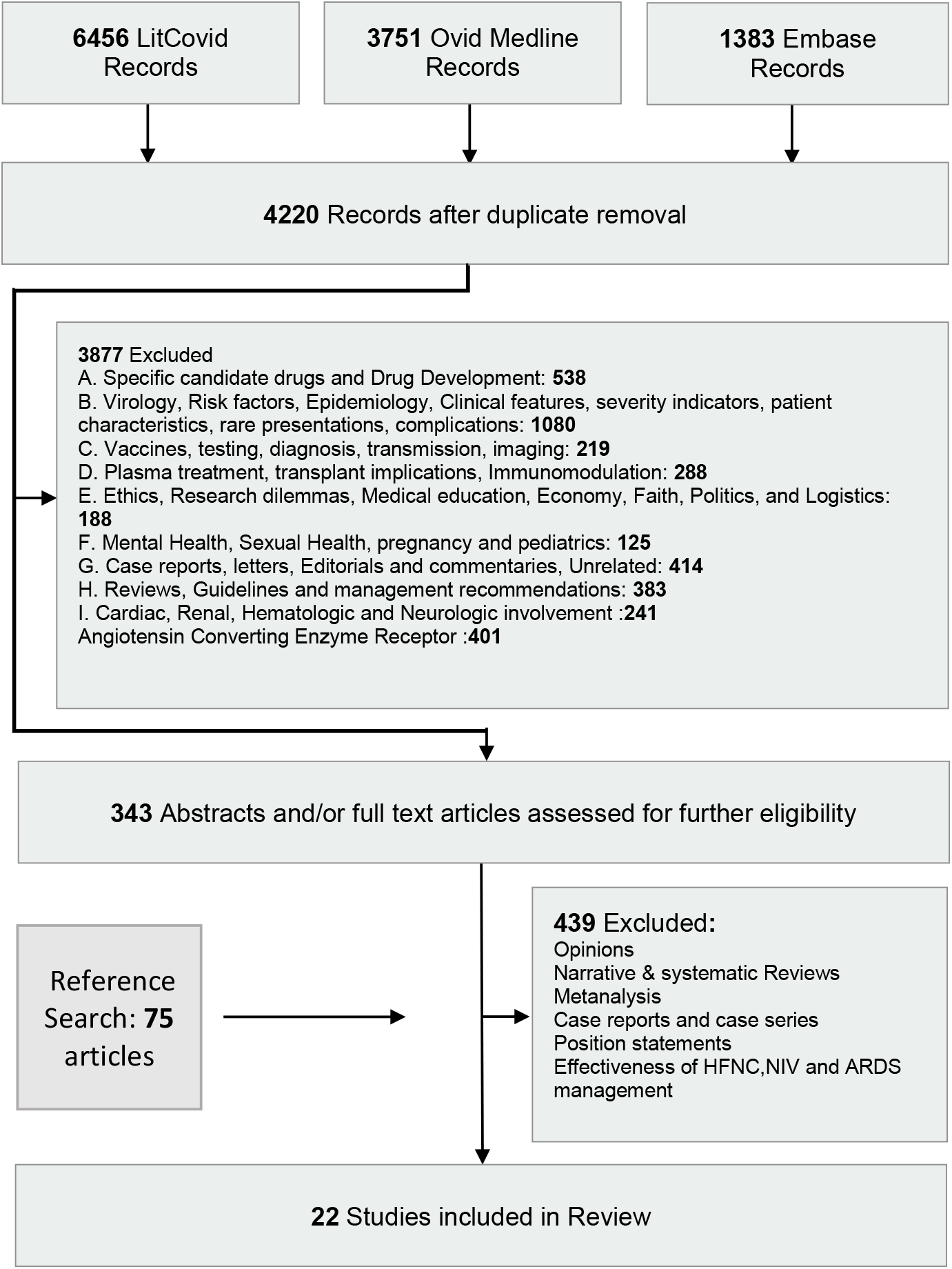

**Table. 1.** Mechanistic studies related to aerosol generation (AGP) and droplet dispersion.

| Study name, Year, Country<br>Setting | Patient/Population<br>Characteristics and<br>Numbers | Intervention or Research question | Outcome | Limitations and<br>Assessment |
| --- | --- | --- | --- | --- |
| **Iwashyna et al 2020 ,USA(18)**<br>Two scanning mobility particle sizing (SMPS) systems (TSI 3080/3030, TSI 3080/3750) were used to measure aerosols 10 to 500 nanometer (nm) in size for each of the oxygen delivery devices(18) | 4 healthy volunteers | 6L/min nasal canula (NC) with humidification.<br>NRB with 15L/min gas flow, non-humidified.<br>HHFNC with 30L/min gas flow.<br>HHFNC with 60L/min gas flow.<br>Measured aerosols (particles from 10 to 500nm in size for each of the oxygen delivery devices<br>Droplet generation measurement only for HFNC.<br>Intermittent forced coughing | There was no variation in aerosol level within patients between room air, 6 L/min NC, 15 L/min NRB, 30 L/min HHFNC, and 60 L/min HHFNC, regardless of coughing. | Small numbers of volunteers, with consequent limitations in the degree of variation in naso/oral structures.<br>None of the volunteers were clinically ill with respiratory condition.<br>CEBM level of evidence 5(mechanistic study)(17) |
| **Kotoda et al 2019,Japan.(25)**<br>Liquid and bacterial dispersal were assessed via in-vitro experimental set-ups using a manikin. Thickened water or fresh yeast solution mimicked saliva and nasal mucus secretions. | Manikin study | Risk of pathogen dispersal during high-flow nasal therapy.<br>HFNC at 60 L·min<sup>-1</sup> compared with HFNC at 0 L·min<sup>-1</sup> | Manual repositioning of the nasal cannula during use of the device slightly increased the dispersal, the dispersal remained limited to the proximal location closest to the manikin's face. No dispersal of water and yeast was detected in areas >60 cm away from the face. | Data only on droplets not on aerosol.<br>Non-Human study<br>CEBM level of evidence 5(Mechanistic study)(17) |
| **Loh et al 2020,Singapore.(37)**<br>Experiment to simulate a patient coughing while using HFNC to assess the maximum distance of droplet dispersion<br>The authors (n = 5), with no history of lung disease, participated | 5 healthy volunteers | Impact of high-flow nasal cannula (HFNC) on coughing distance | Cough-generated droplets spread to a mean (standard deviation) distance of 2.48 (1.03) m at baseline and 2.91 (1.09) m with HFNC. A maximum cough distance of 4.50 m was reported when using HFNC | Properties of food color may not have physical properties of a viral pneumonia patient. Small number of healthy volunteers<br>OCEBM level of evidence 5(mechanistic study)(17) |
| **Leonard et al 2020,USA(26)**<br>Computational fluid dynamic (CFD) simulation to determine the ability of a mask to reduce the velocity of exhaled gas flow and capture particles during HFNC. | Manikin with CFD simulation | HFNC at 40 L·min<sup>-1</sup> compared with nasal prongs at 6 L·min<sup>-1</sup> or spontaneous tidal breathing.<br>Loosely fitted face mask applied to patient simulator | Greater leak (16.5%) with HFNC compared with nasal prongs (12.6%) and spontaneous breathing (11.6%).<br>Droplets captured by face mask were variable with HFNC (85.9%), nasal prongs (75.9%) and spontaneous breathing (89.9%).<br>Variable proportions of escaped particles travelled greater than 1 meter from point of origin with HFNC (15.9%) compared with nasal prongs (6.9%) and spontaneous breathing without mask(31%). | Simulator study<br>Preliminary results<br>CEBM level of evidence level 5(mechanistic study)(17) |
| **Hui, D.S., et al 2006 Hong Kong (24)**<br>studied exhaled air and particle dispersion through an oronasal mask attached to a human-patient simulator (HPS) during noninvasive positive-pressure ventilation (NPPV). Airflow was marked with intrapulmonary smoke for visualization | Manikin with smoke plume visualization | Therapy with inspiratory positive airway pressure (IPAP) was started at 10 cm H<sub>2</sub>O and gradually increased to 18 cm H<sub>2</sub>O, whereas expiratory positive airway pressure was maintained at 4 cm H<sub>2</sub>O. A leakage jet plume was revealed by a laser light sheet and images captured by video. Smoke concentration in the plume was estimated from the light scattered by smoke particles.<br>Experiments were done in proper fit mask setting and loose fit mask setting | A jet plume of air leaked through the mask exhaust holes to a radial distance of 0.25 m from the mask @ IPAP at 10 cm H<sub>2</sub>O with leakage from the nasal bridge most 60 to 80 mm lateral to the median sagittal plane of the HPS.<br>Without leakage, the jet plume from the exhaust holes increased to a 0.40-m radius from the mask, whereas exposure probability was highest about 0.28 m above the patient @IPAP of 10 cm H<sub>2</sub>O. When IPAP was increased to 18 cm H<sub>2</sub>O, the vertical plume extended to 0.45 m above the patient. | Human patient simulator experiment<br>Lung injury setting was not mentioned<br>Limited by using smoke particles as markers for exhaled air which can have significantly different physical property compared to respiratory droplets or aerosols<br>Non comparative.<br>Extrapolation of the result need to be cautious due to significant variations in NIV masks.<br>CEBM level of evidence level 5(mechanistic study)(17) |
| **Hui et al 2009, Hong Kong (23)**<br>Human Patient simulator (HPS) with mild lung injury setting with NPPV support. Intrapulmonary smoke for visualization. A leakage jet plume was revealed by a laser light sheet and captured by high definition video.<br>Isolation room with pressure of -5 Pa. | Manikin study | NPPV with IPAP) started at 10 cm H<sub>2</sub>O and gradually increased to 18 cm H<sub>2</sub>O .<br>EPAP maintained at 4 cm H<sub>2</sub>O .<br>Exhaled air dispersion distances and directions through two different face masks (Respironics; Murrysville, PA) | Substantial exposure to exhaled air occurs within a 1-m region, from patients receiving NPPV via the Comfort Full 2 mask ( 0.65 to 0.85 m) and the Image 3 mask( 0.95 m), compared to Ultra Mirage mask(0.5 m) with more diffuse leakage from the latter, especially at higher IPAP. | Limited by using smoke particles as markers for exhaled air which can have significantly different physical property compared to respiratory droplets or aerosols<br>probably overestimating the dispersion<br>CEBM level of evidence 5(Mechanistic study)(17) |
| **Hui et al 2014, Hong Kong(36)**<br>HPS study as above using high fidelity manikin programmed at different levels of lung injury.<br>Isolation room with pressure of -5 Pa. | Manikin study | Studied the deliberate leakage and dispersion distance from the exhalation ports of Comfort Full 2 and Image 3 masks (Respironics, Murrysville [PA], USA) and other respiratory therapies (jet nebulizer and various oxygen masks) firmly attached to a high-fidelity HPS (Medical Education Technologies, Sarasota [FL], USA). | Non-invasive positive pressure ventilation ResMed Mirage mask at different IPAP/EPAP (cmH<sub>2</sub>O) setting<br>10/4- 0.40-meter ,14/4- 0.42 meter and 18/4 -0.45 meter<br>Respironics Comfort Full 2 mask at different IPAP/EPAP (cmH<sub>2</sub>O) setting<br>10/4 -0.65-meter ,14/4 -0.65 meter and 18/4- 0.85 meter<br>Respironics Image 3 mask plus whisper swivel exhalation valve at different IPAP/EPAP (cmH<sub>2</sub>O) setting 10/4 -0.95 ,14/4 -0.95 and 18/4 >0.95<br>Simple oxygen mask at different oxygen flow in (L/min) 4 -0.20 meter, 6 -0.22 meter, 8 - 0.30 meter ,10 -0.40 and >0.4 during coughing<br>Jet nebulizer (driven by air at 6 L/min) at different lung settings<br>Normal lung 0.45-meter, Mild lung injury 0.54 meter and Severe lung injury >0.80 meter<br>Nasal cannula oxygen (L/min) 1lit -0.30 meter,3lit -0.36 and 5 lit -0.42 meter<br>Venturi oxygen mask Normal lung 24% oxygen- 0.4-meter ,40% oxygen and 0.33 meter<br>Severe lung injury 24% oxygen 0.32 meter and 40% oxygen 0.29 meter<br>Non-rebreathing oxygen mask (oxygen flow, L/min) 6, 8, 10, and 12 <0.1meter | Limited by using smoke particles as markers for exhaled air which can have significantly different physical property compared to respiratory droplets or aerosols<br>probably overestimating the dispersion<br>CEBM level of evidence 5(Mechanistic study)(17) |
| **Hui et al 2015 ,Hong Kong (21)**<br>HPS study in negative pressure room with 12 air changes/h. Air dispersion distance during NIV using Exhaled air was marked by intrapulmonary smoke particles, illuminated by laser light sheet, and captured by a video camera for data analysis. Significant exposure was defined as where there was ≥ 20% of normalized smoke concentration. | Manikin study | Comparison of dispersion distance when providing NPPV via two different helmets and a total facemask via a bilevel positive airway pressure device. | Exhaled air leaked through the neck-helmet interface dispersion distance ranged from 0.15 meter to 0. 27-meter mm when IPAP was increased from 12 to 20 cm H<sub>2</sub>O, respectively, while keeping the expiratory pressure at 10 cm H<sub>2</sub>O.<br>NIV via a helmet with air cushion around the neck, there was negligible air leakage.<br>During NIV via a total facemask for mild lung injury, air leaked through the exhalation port to | Simulated experiment<br>Limited by using smoke particles as markers for exhaled air which can have significantly different physical property compared to respiratory droplets or aerosols with probably |
|  |  |  | 0.618 meter and 0.812meter when inspiratory pressure was increased from 10 to 18 cm H<sub>2</sub>O, | overestimating the dispersion.<br>CEBM level of evidence 5 (mechanistic study)(17) |
| **Hui et al 2019,Hong Kong (22)**<br>HPS was programmed to represent different severity of lung injury. Exhaled airflow was marked with intrapulmonary smoke for visualisation and revealed by laser light-sheet. Normalised exhaled air concentration was estimated from the light scattered by the smoke particles. Significant exposure was defined when there was $\geq 20\%$ normalised smoke concentration. | Manikin study | CPAP was delivered at 5-20 cmH<sub>2</sub>O *via* nasal pillows (Respironics Nuance Pro Gel or ResMed Swift FX) or an oronasal mask (ResMed Quattro Air). HFNC, humidified to 37°C, was delivered at 10-60 L·min<sup>-1</sup> to the HPS. | In the normal lung condition, mean±sd exhaled air dispersion, along the sagittal plane, increased from 186±34 to 264±27 mm and from 207±11 to 332±34 mm when CPAP was increased from 5 to 20 cmH<sub>2</sub>O *via* Respironics and ResMed nasal pillows, respectively. Leakage from the oronasal mask was negligible. Mean±sd exhaled air distances increased from 65±15 to 172±33 mm when HFNC was increased from 10 to 60 L·min<sup>-1</sup>. Air leakage to 620 mm occurred laterally when HFNC and the interface tube became loose. | Used smoke particles as markers of exhaled air.<br>Human Patient Simulator<br>CEBM level of evidence 5 (mechanistic study)(17) |
| **Simonds et al 2010(20)**<br>Three groups were studied: (1) normal controls, (2) subjects with coryzal symptoms and (3) adult patients with chronic lung disease who were admitted to hospital with an infective exacerbation. Each group received oxygen therapy, NIV using a vented mask system and a modified circuit with non-vented mask and exhalation filter, and nebulized saline. The patient group had a period of standardized chest physiotherapy treatment. | 44 subjects and patients were studied: 12 normal controls, 11 with coryzal symptoms and 21 patients of chronic lung disease with exacerbation. | Evaluation of the characteristics of droplet/aerosol dispersion around delivery systems during non-invasive ventilation (NIV), oxygen therapy, nebulizer treatment and chest physiotherapy by measuring droplet size, geographical distribution of droplets, decay in droplets over time after the interventions were discontinued. | NIV using a vented mask produced droplets in the large size range ($> 10 \mu\text{m}$) in patients ($p = 0.042$) and coryzal subjects ($p = 0.044$) compared with baseline values, but not in normal controls ($p = 0.379$), but this increase in large droplets was not seen using the NIV circuit modification. Chest physiotherapy produced droplets predominantly of $> 10 \mu\text{m}$ ($p = 0.003$), which, as with NIV droplet count in the patients, had fallen significantly by 1 m. Oxygen therapy did not increase droplet count in any size range. Nebulized saline delivered droplets in the small- and medium-size aerosol/droplet range but did not increase large-size droplet count. | Used droplets as a proxy for viral dissemination.<br>Chronic lung disease physiology may not accurately represent acute lung injury related to viral pneumonia neither coryzal subjects.<br>CEBM level of evidence 5 (mechanistic study)(17) |
| **Leung et al 2019 ,Honk Kong (27)**<br>Critically ill patients with Gram-negative pneumonia<br>Randomized controlled crossover non-inferiority trial Correlation of Exposure characteristics and development of Probable or suspected SARS | 20 critically ill patients with Gram-negative pneumonia requiring oxygen support (via nasal prongs or face mask) were recruited. | Evaluated the degree of environmental contamination by viable bacteria associated with the (18)use of HFNC 60 l/min compared with conventional oxygen mask. | No difference in GNB count between the HFNC and OM use for air samples, settle plates at 0.4 or 1.5 m, and at six or at 12 air change per hour ($P = 0.119-0.500$). | Study based on bacterial pathogens and gas flow rate used with the OM was relatively high.<br>Small study group<br>Mostly mechanistic study /indirect evidence<br>CEBM level of evidence 5.(17) |

**Table. 2.**
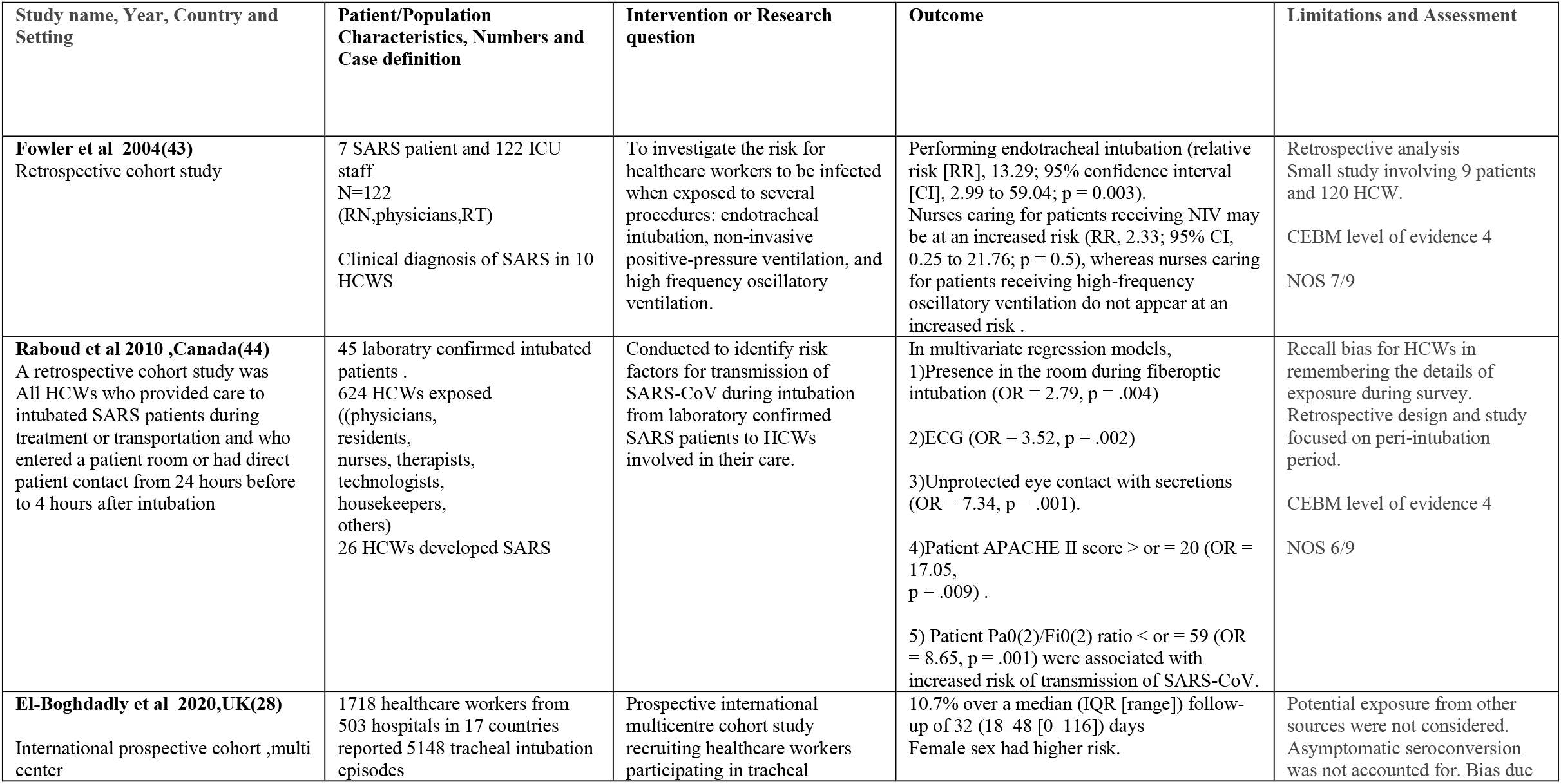
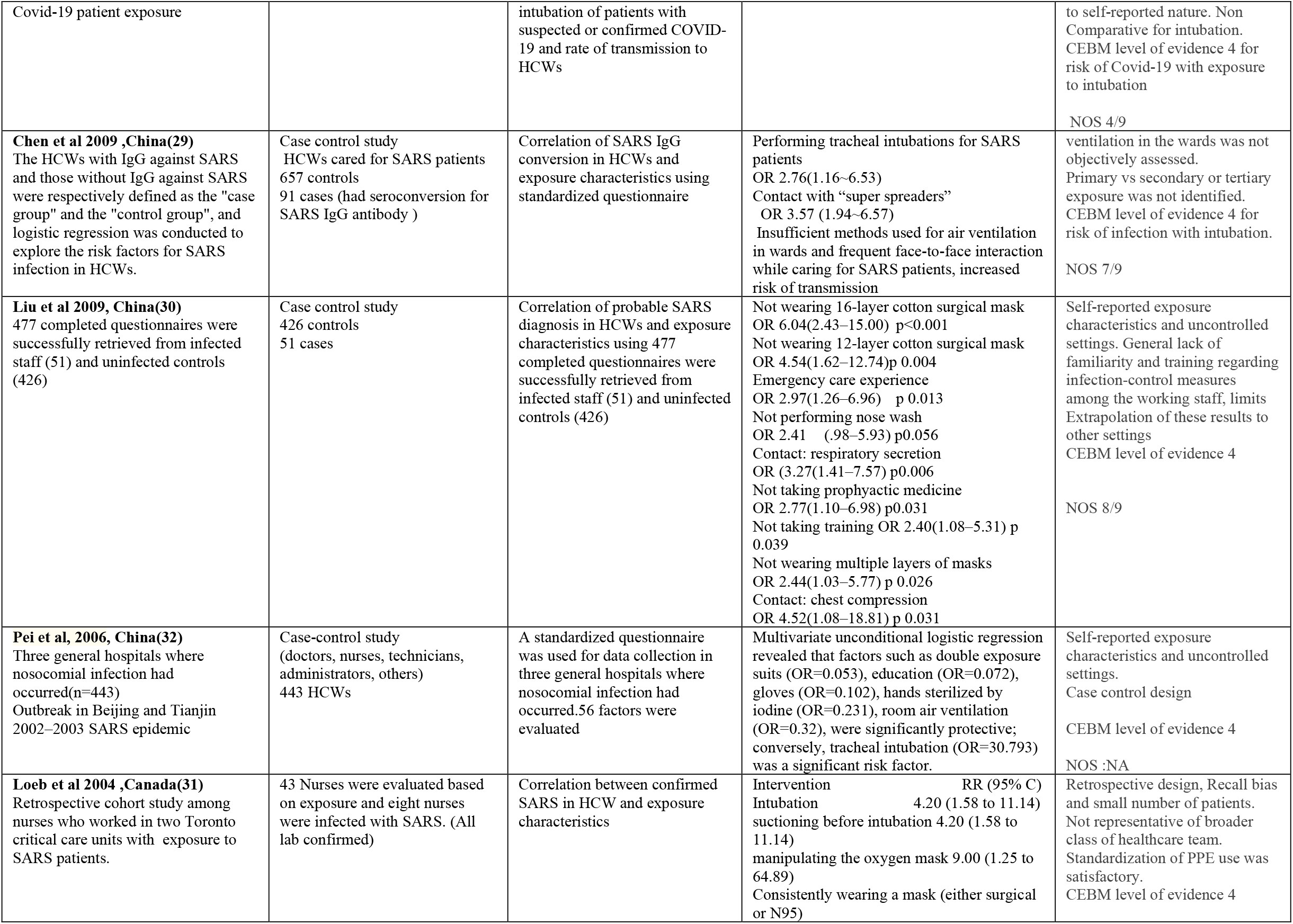
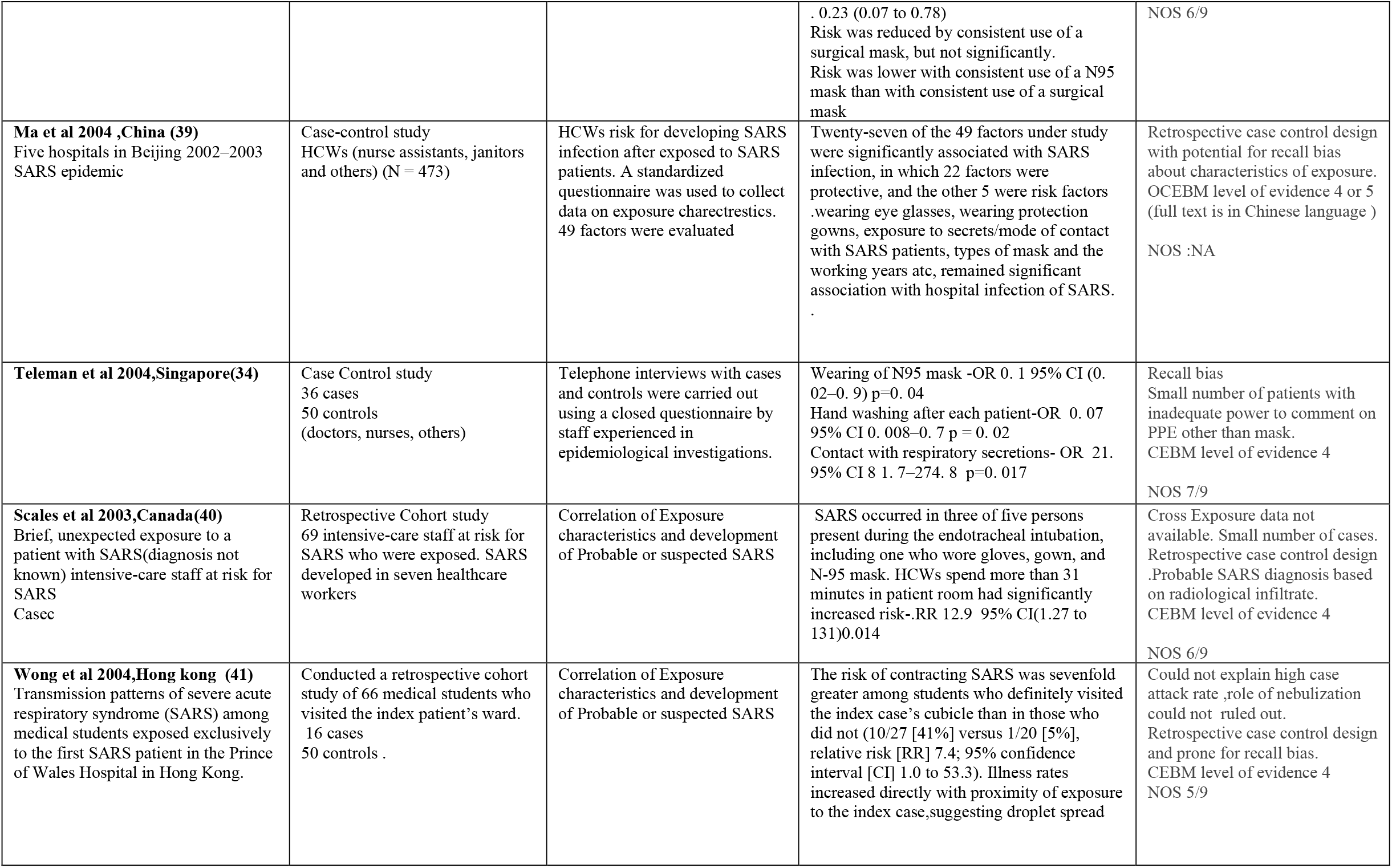
Clinical studies exploring risk of transmission of respiratory infections to HCWs.

## Results

Factors influencing nosocomial transmission of respiratory corona virus infections are HCW dependent factors, Patient characteristics and specific intervention related risks. HCWs dependent factors are related to knowledge, training and compliance with guideline recommended PPE use. Patient characteristics like Acuity of illness, high viral load related “super spreader”, severe respiratory illness and asymptomatic or minimally symptomatic career state also influence HCWs risk of contracting infection. A discussion on the characteristics of super spreaders or evidence based effectiveness of specific type of PPE is beyond the scope this manuscript. In this review, We focused on risk of transmission to frontline HCWs while delivering respiratory supportive care like High flow nasal cannula (HFNC),nasal cannula oxygen, Noninvasive and invasive ventilation, chest physiotherapy and administration of nebulized medication. We also evaluated protective effect of PPE use and patient characteristics only within the context of this supportive care. We found 21 **(Table.1 and Table.2)** studies addressing intervention related exposure risk. Three studies involved healthy volunteers(18-20). Seven out of 2 studies used Human Patient simulator (HPS)(21-26)A prospective randomized crossover design in the context of gram negative bacterial infection was also based on mechanistic of droplet spread(27). One large international prospective cohort was observational in nature.(28)Rest of the studies were retrospective case control or retrospective cohort design(26, 29-34). In total, 11 studies were mechanistic based (**Table.1**) and 11 studies were observational clinical studies **(Table.2)**.

### Mechanistic Studies

Four studies (18, 25-27)compared the risk of HFNC to routine oxygen support or no intervention in terms of aerosol generation or dispersion of respiratory secretion using various surrogates depending on the design. Two of these studies were Human Patient Simulator(HPS) based. Study by Kotoda et al suggested no difference in droplet dispersion between 60 L/Min HFNC in comparison to 0 L/min (no intervention)(25).More recent study using HPS compared HFNC at 40 L·min^− 1^ compared with nasal prongs at 6 L·min^− 1^ and face mask applied to patient simulator looked in to overall droplet dispersion distance, percentage of air leak and effectiveness of facemask loosely applied over oxygen delivery device. Leak was more common for HFNC followed by NC oxygen followed by spontaneous breathing at sites of intentionally created gaps. But percentage of droplets (>5um) cleared by mask was better in HFNC setting. HFNC at 40 L/min captured 83.2% of particles; LFO_2_ at 6 L/min captured 73.6% of particles; and tidal breathing (no therapy) captured 87.2% of particles. Among particles escaped through the gaps in mask almost 16 % in HFNC setting and 7 % in LFO_2_ traveled more than 1-meter distance. But this was 31 % in case of a spontaneous breathing without a mask. (26)The other two studies which compared the effect of oxygen delivery devises to cause particle dispersion, one was involving healthy volunteers (18)and other was in critically ill patients with gram negative pneumonia(27). Human volunteer study did not demonstrate significant difference in aerosol level between room air, 6 L/min NC, 15 L/min NRB, 30 L/min HHFNC, and 60 L/min HHFNC, regardless of coughing. This study focused on extent of aerosol generation than droplet traveling distance. Study on critically ill patients with gram negative pneumonia were based on bacterial dispersion through droplet in four experimental conditions based on method of oxygen support and room ventilation parameters. Use of HFNC or simple oxygen mask (OM) with room ventilation of six or 12 air changes per hour, did not show difference in Gram Negative Bacterial count between HFNC and OM use in air samples, settle plates at 0.4 or 1.5 m, and at six or at 12 air change per hour (*P* = 0.119–0.500).Another study recreated clinical scenario of coughing while using HFNC using healthy volunteers without applying mask on them. Cough-generated droplets spread to a mean (standard deviation) distance of 2.48 (1.03) m at baseline and 2.91 (1.09) m with HFNC. A maximum cough distance of 4.50 m was reported when using HFNC(19).In this particular study, possibility of poorly matched characteristics of liquid substitute used (food colored water)to actual respiratory secretions. None of other studies demonstrated similar results.

Hui et al have done multiple droplet dispersion experiments using HPS in different settings of respiratory support.(21-23, 35).He used intra pulmonary smoke to mark airflow to aid visualization using thin laser light sheet. His models were closely replicative of an isolation room patient. In his 2006 study he explored the extent of dispersion of small particles (< 1 um) while HPS was connected to NIV using a facial mask. In different pressure settings (max IPAP 18 cm of H _2_O) maximum dispersion was less than 0.5 meter(35). When he repeated this experiment in 2009 using two different facemasks, dispersion was up to 1-meter distance and there was difference in dispersion in relation to mask exhaust characteristics(23). His similar experiment in 2015 which tested two different helmet interface and a total facemask connected to a bilevel positive airway pressure device with 12 air changes/h in the room demonstrated significant air dispersion with facemask exhalation port to 618 and 812 mm when inspiratory pressure was increased from 10 to 18 cm H2O, respectively, with EPAP at 5 cm H2O, but dispersion through the neck-helmet interface with a radial distance of 150 to 230 mm when IPAP was increased from 12 to 20 cm H2O, respectively. During NIV via a helmet with air cushion around the neck, there was negligible air leakage(21).Similar experiment involving 3 types of subjects 1)healthy volunteers 2)patients with simple coryza or 3)exacerbation of chronic lung disease, evaluated the characteristics of droplet/aerosol dispersion by measuring droplet size, geographical distribution of droplets, decay in droplets over time, and the impact of modification of the NIV circuit. This study evaluated various respiratory care procedures.(NIV, O2, nebulizer treatment or chest physiotherapy). NIV using a vented mask caused increase production of droplets in the large size range (> 10 μm)in patients (*p* = 0.042) and coryzal subjects(*p* = 0.044) compared with baseline values, but not in normal controls (*p* = 0.379). This increase in large droplets did not happen when NIV circuit was modified with non-vented mask and exhalation filter. Chest physiotherapy increase production of droplets predominantly of > 10 μm (*p* = 0.003), which, as with NIV droplet count in the patients, had fallen significantly by 1 meter. O2 did not increase droplet count in any size range. Nebulization increased in the number of small- and medium-size aerosol/droplet range but did not increase large-size droplet count. Preliminary analysis suggested that droplet counts fall to within a baseline range in 20–40 minutes of discontinuing the NIV and chest physiotherapy(20).Another experimental study by Hui et al compared effect of CPAP with different types of patient interfaces or HFNC and simulated different lung conditions using programmable HPS. In normal lung condition, the exhaled air dispersion distances from HFNC along the median sagittal plane increased significantly with increasing flow rate from 10 L·min− 1 60 L·min− 1 from to a mean±SD of 65±15mm to 172±33 mm (p<0.001).In an interesting note when lung condition was programmed to severe lung injury, this distance fell to 48±16 mm.(22).Hui et al did more comprehensive simulation study in 2014 which looked at the effect of deliberate leakage and dispersion distance from the exhalation ports of Comfort Full 2 and Image 3 masks (Respironics, Murrysville [PA], USA) and other respiratory therapies (jet nebulizer and various oxygen delivery devices) using a high-fidelity HPS. When tested ResMed Mirage mask, Respironics Comfort Full 2 mask and Respironics Image 3 mask plus whisper swivel exhalation valve, at higher setting (IPAP 18 cm of H_2_O) dispersion distance varied from. 45 meter to 1 meter lowest being for ResMed Mirage mask and highest for the one with swivel exhalation valve. Comparative dispersion rate for nasal cannula oxygen at 5-6 litter/mt was significantly higher than simple oxygen mask (0.22 meter vs 0.42 meter).A jet nebulizer driven by 6 lit/mt oxygen flow had a dispersion distance up to 1 meter when manikin setting was for severe lung injury. Venturi oxygen mask in severe lung injury setting had 0.33 meter dispersion, but Non-rebreathing oxygen mask had very low dispersion distance (Table.1)(36)

### Clinical Studies

10 retrospective observational studies (5 case control and 5 retrospective cohort) and one prospective observational study explored of procedures and behaviors modifies risk of transmission corona virus infection to HCWs during routine respiratory supportive care **(Table.2)**. All except one (28)of these studies were done in the context of SARS-Cov-1 infection(28-34, 38-40).Study by Fowler et al which specifically looked into factors like exposure to intubation procedure, NIV or HFNC treatment modalities. His study clearly showed strong association to exposure to intubation for transmission of SARS infection to HCWs. But caring for patients with invasive ventilation did not increase the risk of infection to frontline providers. Nurses caring for patients receiving noninvasive positive-pressure ventilation showed a trend towards increased risk (RR, 2.33; 95% CI, 0.25 to 21.76; p = 0.5) but did not reach statistical significance(38).8 out of 9 studies consistently showed significantly increased risk of Corona virus infection to HCWs if present during intubation related procedures **(Table.2)**.One of this study worth special mention is a recent large multinational prospective registry study specifically focused on intubation related risk of exposure to SARS-Cov-2 virus infection in HCWs, which demonstrated a whopping 11 % incidence of transmission of Covid-19 to healthcare providers(28). Only one of the three studies looked at the impact of emergency care setting suggested increased risk of infection to HCWs(30).But two of three studies looked at the effect of exposure time during individual visit by HCW, suggested increased risk of nosocomial infection(32, 41).6 out of 7 studies favored mask use, preferably multilayered or surgical mask(4, 8, 28, 29, 31, 36, 39). Four of five studies did show benefit of universal use of N 95 mask in comparison to surgical mask(30, 31, 34, 39-41). Out of eight studies looked at effect of glove use, five showed significant reduction in transmission of infection(28-34, 40). But six out of nine studies showed preventive potential of wearing a gown(28-30, 32-34, 39, 40).**(Table.2)**.Two of the three studies looked at the effect of post exposure handwashing showed significant decrease in transmission of infection and third study showed similar trend without reaching statistical significance (29, 32, 34). Similarly, all four studies looked at the effect of eye protection showed robust decrease in transmission(29, 30, 33, 39). All three studies explored the risk of contact with respiratory secretion showed increased risk of contracting infection(30, 34, 39). When two studies looked at the correlation of severity of respiratory disease in index patient to infectious potential, both demonstrated positive correlation for transmissibility to HCWs. Potentially such patients may be immunocompromised with high viral load and become ‘super spreaders(30, 33). Another significant factor emerged as protective based on evidence was formal infection control training to HCWs. All four studies looked at this concurred to support formal training(29, 30, 32, 33)(Table.2).Risk of infection spread in the setting Noninvasive ventilatory support was explored by two studies. Both study showed trend towards increased transmissibility of SARS to HCWs in univariate analysis, but did not maintain statistical significance on multivariate regression(33, 38). On a similar note, effect of HFNC support in nosocomial spread of Corona virus infection was evaluated by two studies(Raboud et al-SARS-Cov-1, El-Boghdadly et al-SARS-Cov-2).Neither of them showed statistically significant correlation of HFNC use and infection rate in HCWs(28, 33)

### Quality and strength of Evidence-Mechanistic studies

Mechanistic studies are a great way of addressing clinical questions especially safety of a particular intervention is considered. Simulation can offer researchers access to events that can otherwise not be directly observed, and in a safe and controlled environment. It allows for controlled variation of variable. In the same note simulation or other mechanistic studies are considered indirect and hypothetical evidence. Most studies we reported in this summary had reasonable fidelity, context appropriateness with good quality of reporting. None of these study designs were externally validated(42).Studies looked at aerosol generation potential of interventions were very few and only for very limited interventions(15, 18, 35)One study strongly suggested nebulization is associated with significant increase in aerosol range particles. Aerosol generation potential for HFNC, NIV, chest physiotherapy or other oxygen delivery methods were significantly low. Placing a well-fitting facemask above patient interface almost nullified both aerosol and droplets escaping from the circuit (Table.1). Five studies looked at droplet dispersion potential of HFNC using healthy volunteers, bacterial pneumonia patients, patients with coryza or Chronic lung disease(22, 25-27, 37). In the setting of a well fitted nasal interface, dispersion ranged between 0.18 meter to less than 0.5 meter and most instance in the range of 0.2 to 0.3-meter distance. There was some degree of association with oxygen flow rate to dispersion distance. Dispersion distance for HFNC was not significantly different than Nasal cannula oxygen. Placing a well fitted face mask reduced droplets by more than 85%. This were true even during coughing episodes except in 1 study with questionable fidelity(37).When six studies looked at the droplet dispersion potential of NIV, there were significant difference in the results(20-23, 35, 36).Studies consistently showed that dispersion distance is highly dependent on the type of patient interface(modified vs exhalation valves on the front of mask).With modified NIV circuit or helmet interface with neck cushions, dispersion was almost nonexistent. Dispersion distance could be decreased to the level of nasal cannula oxygen when modified circuitry employed **(Table.1)**.Major drawback of simulation studies are, they are inherently very low quality evidence when extrapolating to real clinical scenario and level of evidence is usually falls in the range of 4 to 5 at best.Other major limitation of such experimental studies is inability to draw a conclusion on effect size or impact.

### Quality and strength of Evidence-Clinical Studies

We used Newcastle - Ottawa quality assessment Scale (NOS-Appendix 4)(16) for observational studies to evaluate clinical studies. Five of the clinical studies were case control design. NOS employs ‘star system’ in which a study is judged on three broad perspectives 1)the selection of the study groups 2)the comparability of the groups and 3)the ascertainment of either the exposure or outcome of interest for case-control or cohort studies respectively. Among case control studies, We could not evaluate NOS for 2 studies with Chinese language full text.(32, 39). NOS score of Two studies were 7/9 (29, 34)and one study was 8/9(30) (Appendix 4).One of the six cohort study was prospective design(28) and rest were retrospective design(31, 33, 38, 40, 41). NOS score was 7/9 for one study(33),6/9 for three studies (31, 38, 40) 5/9 for one study(41) and 4/9 for one study(28).Details of evaluation tool is available as supplementary table(Appendix 3).Observative nature of studies make most of them low quality studies with CEBM level of evidence ranging from 3a to 5 and most instances it was level 4 (Table.2).Grade of evidence for risk or benefit of interventions incorporating CEBM level of evidence(17) and NOS score(16) for individual studies gathered from this review is given below(Table.3).When considering GRADE of evidence, we rated up or down depending on factors like effect size, NOS score(risk of bias) and consistency of results from different studies. Grading of Recommendations, Assessment, Development and evaluations (GRADE) scoring was followed to report strength and quality of evidence for each interventions(Appendix 4)(45).We did not attempt to pool data to quantitatively asses effect size due to highly variable designs and settings of the study. Different studies approached same question with different approach. For example, Loeb et al and Teleman et al addressed role of PPE as effect of commission whereas as Chen et al looked at the effect of omission**(Table.3)**. Similarly multiple designs of the study (case control, retrospective cohort and prospective cohort) was a major barrier for quantitative pooling.

**Table. 3.** GRADE (Grading of Recommendations Assessment, Development and Evaluation) for interventions(17)

| **Intervention** | **Increased Risk (RR/OR )** | **Decreased Risk (RR/OR)** | **No difference (RR/OR)** | **GRADE** |
| --- | --- | --- | --- | --- |
| Intubation | Liu et al (NA)<br>Fowler et al 0.74(0.11 to 4.92)<br>Raboud et al 2.79 (1.40-5.58),<br>Loeb et al 4.20 (1.58 to 11.14)<br>Pie et al (OR=30.793) ,<br>Scales et al (NA)<br>El-Boghdadly et al (NA) ,<br>Chen et al 8.03 (3.90~16.56) |  | Teleman et al 1. 5 (0. 4–5. 4) p=0. 6 | Moderate certainty for harm<br>GRADE B evidence |
| CPR/emergency care | Liu et al 4.52(1.08–18.81) p 0.031 |  | Raboud et al (NA)<br>El-Boghdadly et al (NA) | Very low certainty evidence<br>for harm<br>GRADE D |
| Prolonged Exposure during individual visit | Wong et al 7.4(1.0 to 53.3)<br>Scales et al 12.9(1.27 to 131) |  | Teleman et al 0. 7 (0. 3–1. 6) 0. 4 | Low certainty evidence for harm<br>GRADE C |
| Any Mask |  | Liu et al 2.44(1.03–5.77) p0.026<br>Raboud et al (NA)<br>Ma et al (NA)<br>Scales et al (NA)<br>Loeb et al 0.23 (0.07 to 0.78)<br>Chen et al 2.53 (1.57~4.07). | El-Boghdadly et al (NA) | Moderate certainty evidence<br>for benefit<br>GRADE B |
| N 95 Mask compared to regular mask in routine patient care |  | Ma et al (NA)<br>Scales et al (NA)<br>Teleman et al 0. 1 (0. 02–0. 9)<br>Loeb et al 0.22 (0.05 to 0.93) p 0.06 | Liu et al (NA) | Low certainty evidence for benefit over regular mask<br>GRADE C |
| Glove |  | Liu et al (NA)<br>Scales et al (NA)<br>Pie et al (OR=0.102)<br>Chen et al 5.20 (2.65~10.23)<br>Teleman et al, 0. 1 (0. 03–0. 4) | Raboud et al(NA)<br>El-Boghdadly et al (NA)<br>Loeb et al 0.45 (0.14 to 1.46) | Low certainty evidence for benefit<br>GRADE C |
| Gown |  | Liu et al(NA)<br>Raboud et al(NA)<br>Ma et al(NA),<br>Pie et al (OR=0.053)<br>Scales et al(NA),<br>Chen et al 2.12 (1.36~3.31) | Loeb et al 0.36 (0.10 to 1.24)<br>El-Boghdadly et al (NA)<br>Teleman et al 0. 5 (0. 1–1. 4) | Moderate certainty evidence for benefit<br>GRADE B |
| Post exposure hand washing |  | Pie et al (OR=0.231)<br>Teleman et al 0. 06 (0. 007–0. 5) | Chen et al 0.89 (0.52~1.51) >0.05 | Low certainty for benefit<br>GRADE C |
| Post Exposure nasal rinsing |  | Liu et al,2.41(0.98–5.9) p 0.056 | Chen et al 3.21 (0.98~10.53) >0.05 | Very low certainty evidence for benefit<br>GRADE D |
| Prophylactic Medicine |  | Liu et al 2.77(1.10–6.98) p0.031 |  | Very low certainty evidence for benefit<br>GRADE D |
| Goggle |  | Liu et al(NA)<br>Ma et al (NA),<br>Chen et al 7.83 (1.07~57.63)<br>Raboud et al 7.34(2.19-24.52) |  | Moderate certainty evidence for benefit<br>GRADE B |
| Contact with respiratory secretions | Liu et al 3.27(1.41–7.57),<br>Teleman et al 6. 9 (1. 4–34. 6)<br>Ma et al(NA), |  |  | Moderate certainty evidence for harm<br>GRADE B |
| Contact with other secretions |  |  | Liu et al (NA) | Very low certainty evidence harm<br>GRADE D |
| Severe disease/lung injury | Liu et al( NA),<br>Raboud et al 8.65 (2.31-32.36) |  |  | Low certainty evidence for harm<br>GRADE C |
| Formal Infection Control training |  | Liu et al 2.40(1.08–5.31)<br>Pie et al (OR=0.072)<br>Raboud et al(NA),<br>Chen et al 2.44 (1.41~4.23) |  | Moderate certainty evidence for benefit<br>GRADE B |
| Patients on NIV | Raboud et al (NA) |  | Fowler et al 2.33(0.25 to 21.76) p 0.5 | Very low certainty evidence for harm<br>GRADE D |
| Patients on HFO |  |  | Fowler et al 0.74(0.11 to 4.92) p 0.6 | Very low certainty evidence for Harm<br>GRADE D |
| Patients on HFNC | El-Boghdadly et al (NA) |  | Raboud et al(NA) | Very low certainty evidence for Harm<br>GRADE D |
| Invasive mechanical ventilation |  |  | Raboud et al(NA) | Very low certainty evidence for Harm.<br>GRADE D |
| Nebulizer treatment | Loeb et al 3.24 (1.11 to 9.42) p 0.09 |  | Raboud et al (NA) | low certainty evidence for harm<br>GRADE C |
| Number of shifts worked | Loeb et al(NA) |  | Fowler et al(NA) | Very low certainty evidence for harm<br>GRADE D |
| Appropriate Doffing technique |  |  | Raboud et al (NA) | Very low certainty for benefit<br>GRADE D |
| Suctioning after intubation | Loeb et al 0.68 (0.21 to 2.26) 0.04 |  | Raboud et al(NA), Teleman et al (NA) | Very low certainty for harm<br>GRADE D |
| Suctioning before intubation | Loeb et al 4.20 (1.58 to 11.14) |  | Raboud et al(NA), Teleman et al(NA) | Very low certainty for harm<br>GRADE D |
| Bag and mask ventilation | Raboud et al(NA) |  | El-Boghdadly et al 0.81 (0.54, 1.23) | Very low certainty for harm or no harm<br>GRADE D |
| Manipulation of oxygen mask | Raboud et al(NA)<br>Loeb et al 9.00 (1.25 to 64.89) |  |  | Low certainty for Harm<br>GRADE C |
| Decreased Face to face contact | Chen et al 0.16 (0.06~0.46) <0.001 |  | Raboud et al(NA) | Very low certainty for harm or no harm<br>GRADE D |
| Number of times entered in patient room |  |  | Raboud et al(NA) | Very low certainty for harm or no harm<br>GRADE D |

Overall Among the intervention putting HCWs at risk of infections, intubation consistently proven to be a high-risk procedure with moderate certainty(GRADE B). Evidence gathered from this review support use of NIV with modified circuit or helmet interface with sealing cushions at neck body. Evidence against NIV regarding risk of transmission were based on two retrospective studies with very low certainty evidence(33, 38) and results were not statistically significant. Two non-comparative reports published based on Covid-19 exposure did not show significant risk, but primary outcome happened only in 3 HCWs in one study and none in the other (46, 47) so we did not include these studies for the purpose of review. Risk of transmission with Emergency resuscitation measures based on very low certainty evidence (GRADE D). Exposure time and number of shifts also showed to be correlating with risk of transmission based on low certainty evidence to HCWs(GRADE C)(32, 34). Similarly when studies looked at tracheal suction after and before intubation only one study suggested an increased risk in both setting(very low certainty, Grade D) (31, 33, 34). Evidence against safety of nebulizer treatment from clinical studies were contradictory and did not reach statistical significance(Very low certainty, GRADE D)(31, 33).Similarly evidence to support risk of transmission with Bag and Mask ventilation was extremely weak and of very low certainty. Studies looked at the risk of transmission with HFNC, did not show significant correlation and finding was of two opposing trend (28, 33).On similar note two studies looked at effect of caring for mechanically ventilated patients. There was no increased trend for transmission risk(33, 38).Potentially there is stronger evidence for transmission risk with manipulation of patient interface (low certainty, GRADE C)(31, 33).Contact with respiratory secretion also showed similar trend(GRADE C) **(Table.3)**(30, 34, 39).Discussion regarding infection prevention in this setting was not a focus of our study, but data from above studies clearly showed benefit of any multilayered mask or surgical mask(Moderate certainty-GRADE B)(29-31, 33, 34, 39, 40).Five studies looked at benefit of N 95 mask in comparison to regular surgical mask. There is low certainty evidence (GRADE C) to support the use. Similarly evidence supporting use of gloves were also low certainty (GRADE C)**(Table.3)**. There is moderate certainty evidence to support use of multilayered clothing or gown to prevent transmission of corona virus to HCWs(GRADE B)(3, 7, 27, 28, 30, 34, 35, 38).On similar note Formal training provided to HCWs on PPE had significant impact on decreasing transmission(GRADE B)(29, 30, 32, 33).Another intervention with significant impact on preventing transmission was wearing eye protection. All four studies reported statistically significant benefit of wearing goggles or eye shields(29, 30, 33, 39)(GRADE B).

## Discussion

Our literature review for identifying factors and procedures involved in respiratory supportive care of patients with corona virus pneumonia identified 21 studies based on our inclusion(Figure.1).Till date there is Despite being indirect and observational studies, we could draw some important conclusions to improve safety of HCWs delivering care to these patients. One of the strengths of our review is combining mechanistic and clinical studies. Nevertheless mechanistic studies cannot make an assessment of effect size, valuable information can be used to design clinical studies and support or dispute low or very low certainty evidence generated from clinical studies. Available evidence to guide frontline health care providers to safely deliver respiratory care to Covid-19 or other corona virus pneumonia is very limited. Few systematic reviews from the past attempted to address this in the context of SARS.(7)Tran et al did not include mechanistic studies in this review and risk of bias assessment of individual studies were not performed. Significant risk factors identified in this review were tracheal intubation, non-invasive ventilation, tracheotomy and manual ventilation before intubation.. We also found performing or assisting endotracheal intubation have significant risk. Preliminary result from recent multinational prospective observational study suggested almost 11 % incidence of Covid-19 in HCWs performing Tracheal intubation. Result of this uncontrolled study need to be interpreted with caution(28). Despite that healthcare system logistics need to be mindful of the potential immediate impact on availability of specialized human resource not to mention the emotional and physical health of the HCWs. In some instances, even with recommended PPE some providers got infected. More methodologically sound prospective observational studies needed to identify risk mitigation strategies for HCWs during this life saving procedure for the patient. Multiple experimental interventions being tried including protective membranes or ‘box’. Proper use of PPE including eye protection and frequent hand hygiene remains most important factors with strong evidence to mitigate transmission to HCWs. (48)

Major confusion among frontline health care providers at the onset of Covid-19 pandemic was about safety of Nebulization, HFNC and NIV support to Covid-19 patients. Being in the midst of a pandemic these are very important questions needing immediate answers. Unfortunately quality studies on Covid-19 addressing these issues are yet to be published. Tran et al did not find significant risk of transmission with nebulization treatment. In our review one study showed possible trend for increased transmission(31)but did not reach statistical significance(p value0.09,NOS 6/9). On multivariate regression Raboud et al did not find nebulization as a risk factor(NOS 7/9)(33).But two mechanistic studies suggested increased aerosol generation and increased droplet dispersion of up to 1 meter with use of jet nebulizer.(20, 36).This discordance may be reflective of possibility of ineffective dispersion of actual viral particle by aerosol created by nebulization. Until we have more evidence from randomized studies or great quality observational study, it is better to avoid nebulization in Covid-19 patients. Similar dilemma also exists in the use of NIV and HFNC in the setting of corona virus pneumonia. Recent metanalysis of noninvasive oxygenation modalities(NIV,HFNC and Oxygen treatment) had shown RR of 0.26, 0.76 and 0.76 for Helmet noninvasive ventilation, face mask noninvasive ventilation and high-flow nasal oxygen respectively in comparison to conventional oxygen treatment for endotracheal intubation. Both NIV modalities also significantly decreased mortality(49)In this review both studies looked at risk of NIV to cause nosocomial viral Pneumonia did not show statistically significant effect on multivariate analysis(33, 38).Mechanistic studies looked at impact of NIV showed highly variable result depending on the patient interface and exhalation circuit. Dispersion of droplets were insignificant with helmet interface and modified exhalation circuit **(Table.1)**.One clinical study which suggested possible increase in transmission with HFNC (3% vs 6%) is based on unadjusted raw data (28) and has a low NOS score(4/9) in comparison to the study rejected this hypothesis(33)**(Table.2)**.Two mechanistic study involving healthy volunteers had significantly contrasting results.(18, 37)But study design and fidelity of the study set up was significantly compromised in the study which suggested increased risk with HFNC.(37)Manikin based dispersion studies did not show significant difference from nasal cannula oxygen when well fitted nasal interface is used**(Table.1)**.Result was similar when HFNC was tested on Gram negative bacterial pneumonia patients with culture plate placed at different distance(27). Other significant risk factors based on our review were direct contact with respiratory secretion, lack of formal infection control training, not consistently wearing mask, gown or eye protection(GRADE B) **(Table.3)**

Our review has limitations. Due to lack of qualifying comparative studies on Covid-19 supportive care and risk of transmission, this study is not strictly a systematic review. We attempted to extrapolate indirect evidence form previous corona virus epidemics and in some case from non-viral pneumonia.(37) The included original studies had poor reporting quality and were mostly observational. Included mechanistic studies could not draw meaningful conclusion on effect size. Different designs and lack of uniformity in approaching clinical questions deterred a possibility of quantitative pooling of the data without serious risk of bias. Across the spectrum of studies serious confounding due to potential variations in PPE use were not always adjusted.

In conclusion we attempted to answer fundamental concerns of frontline HCWs fighting the Covid-19 pandemic regarding nosocomial infection risk by critically appraising limited available evidence mostly extrapolating from past corona virus epidemics with few additions from current pandemic. We adhered to CEBM level of evidence tool and NOS scoring system to appraise individual studies and incorporated information from mechanistic and volunteer studies while making GRADE assessment of intervention of interest. We found non invasive oxygenation strategies like HFNC, NIV with modified circuit or helmet interface is as safe as nasal cannula oxygen. Endotracheal intubation remains high risk for nosocomial transmission in corona virus and early invasive ventilation strategies not supported by available data. Properly designed prospective studies to identify safe intubation practices is need of the hour to protect HCWs from infection. Even though evidence against risk of transmission with nebulization is low or very low certainty, recommendation is to avoid this intervention till we have more data regarding both safety and efficacy.

## Data Availability

All data referred here are based on review of already peer-reviewed journal publications. Please refer to references for details.

## Acknowledgments

Access to data base and journal search: Carpenter Library

## Conflict of Interest Disclosures

None

## Funding/Support

None

Design and conduct of the Review: TK

Search, collection, management, analysis, and interpretation of the articles:TK,TT,AP Assessment of level and GRADE of evidence:TK

Preparation, review, or approval of the manuscript: All authors Decision to submit the manuscript for publication: All authors

## Primary Funding Source

None

## Disclosure

None of the authors have any conflict of interest to disclose.

## Notes

### Competing Interest Statement

The authors have declared no competing interest.

### Clinical Trial

NA

### Author Declarations

This is a review. No IRB clearance required. No active interventions performed or identifiable data published.No direct raw data analysis performed.

## References

1. Cui J, Li F, Shi Z-L. Origin and evolution of pathogenic coronaviruses. Nature Reviews Microbiology. 2019;17(3):181–92.

2. Wu F, Zhao S, Yu B, Chen Y-M, Wang W, Song Z-G, et al. A new coronavirus associated with human respiratory disease in China. Nature. 2020;579(7798):265–9.

3. de Wit E, van Doremalen N, Falzarano D, Munster VJ. SARS and MERS: recent insights into emerging coronaviruses. Nature reviews Microbiology. 2016;14(8):523–34.

4. Huang C, Wang Y, Li X, Ren L, Zhao J, Hu Y, et al. Clinical features of patients infected with 2019 novel coronavirus in Wuhan, China. Lancet (London, England). 2020;395(10223):497–506.

5. Wang D, Hu B, Hu C, Zhu F, Liu X, Zhang J, et al. Clinical Characteristics of 138 Hospitalized Patients With 2019 Novel Coronavirus– Infected Pneumonia in Wuhan, China. JAMA. 2020;323(11):1061–9.

6. Yang X, Yu Y, Xu J, Shu H, Xia Ja, Liu H, et al. Clinical course and outcomes of critically ill patients with SARS-CoV-2 pneumonia in Wuhan, China: a single-centered, retrospective, observational study. The Lancet Respiratory Medicine. 2020;8(5):475–81.

7. Tran K, Cimon K, Severn M, Pessoa-Silva CL, Conly J. Aerosol generating procedures and risk of transmission of acute respiratory infections to healthcare workers: a systematic review. PloS one. 2012;7(4):e35797–e.

8. Lee N, Hui D, Wu A, Chan P, Cameron P, Joynt GM, et al. A major outbreak of severe acute respiratory syndrome in Hong Kong. N Engl J Med. 2003;348(20):1986–94.

9. Chan-Yeung M, Yu WC. Outbreak of severe acute respiratory syndrome in Hong Kong Special Administrative Region: case report. BMJ (Clinical research ed). 2003;326(7394):850–2.

10. Feng D, de Vlas SJ, Fang LQ, Han XN, Zhao WJ, Sheng S, et al. The SARS epidemic in mainland China: bringing together all epidemiological data. Trop Med Int Health. 2009;14 Suppl 1(Suppl 1):4–13.

11. Reynolds MG, Anh BH, Thu VH, Montgomery JM, Bausch DG, Shah JJ, et al. Factors associated with nosocomial SARS-CoV transmission among healthcare workers in Hanoi, Vietnam, 2003. BMC Public Health. 2006;6:207.

12. Covid C. Characteristics of health care personnel with COVID-19—United States, February 12–April 9, 2020. https://wwwcdcgov/mmwr/volumes/69/wr/pdfs/mm6915e6-Hpdf. 2020.

13. Higgins JP, Thomas J, Chandler J, Cumpston M, Li T, Page MJ, et al. Cochrane handbook for systematic reviews of interventions: John Wiley & Sons; 2019.

14. Chen Q, Allot A, Lu Z. Keep up with the latest coronavirus research. Nature. 2020;579(7798):193.

15. Moher D, Liberati A, Tetzlaff J, Altman DG, Altman D, Antes G, et al. Preferred reporting items for systematic reviews and meta-analyses: the PRISMA statement (Chinese edition). Journal of Chinese Integrative Medicine. 2009;7(9):889–96.

16. Wells G, Shea B, O’Connell D, Peterson j, Welch V, Losos M, et al. The Newcastle–Ottawa Scale (NOS) for Assessing the Quality of Non-Randomized Studies in Meta-Analysis. ?. 2000;?.

17. Medicine OCfE-B. OCEBM Levels of Evidence Working Group. The Oxford 2011 levels of evidence. 2011.

18. Iwashyna TJ, Boehman A, Capelcelatro J, Cohn AM, Cooke JM, Costa DK, et al. Variation in Aerosol Production Across Oxygen Delivery Devices in Spontaneously Breathing Human Subjects. medRxiv. 2020:2020.04.15.20066688.

19. Loh N-HW, Tan Y, Taculod J, Gorospe B, Teope AS, Somani J, et al. The impact of high-flow nasal cannula (HFNC) on coughing distance: implications on its use during the novel coronavirus disease outbreak. Canadian journal of anaesthesia = Journal canadien d’anesthesie. 2020;67(7):893–4.

20. Simonds AK, Hanak A, Chatwin M, Morrell M, Hall A, Parker KH, et al. Evaluation of droplet dispersion during non-invasive ventilation, oxygen therapy, nebuliser treatment and chest physiotherapy in clinical practice: implications for management of pandemic influenza and other airborne infections. Health Technol Assess. 2010;14(46):131–72.

21. Hui DS, Chow BK, Lo T, Ng SS, Ko FW, Gin T, et al. Exhaled air dispersion during noninvasive ventilation via helmets and a total facemask. Chest. 2015;147(5):1336–43.

22. Hui DS, Chow BK, Lo T, Tsang OTY, Ko FW, Ng SS, et al. Exhaled air dispersion during high flow nasal cannula therapy <em>versus</em> CPAP <em>via</em> different masks. European Respiratory Journal. 2019:1802339.

23. Hui DS, Chow BK, Ng SS, Chu LCY, Hall SD, Gin T, et al. Exhaled air dispersion distances during noninvasive ventilation via different Respironics face masks. Chest. 2009;136(4):998–1005.

24. Hui DS, Hall SD, Chan MTV, Chow BK, Tsou JY, Joynt GM, et al. Noninvasive positive-pressure ventilation: An experimental model to assess air and particle dispersion. Chest. 2006;130(3):730–40.

25. Kotoda M, Hishiyama S, Mitsui K, Tanikawa T, Morikawa S, Takamino A, et al. Assessment of the potential for pathogen dispersal during high-flow nasal therapy. J Hosp Infect. 2020;104(4):534–7.

26. Leonard S, Atwood CW, Jr., Walsh BK, DeBellis RJ, Dungan GC, Strasser W, et al. Preliminary Findings on Control of Dispersion of Aerosols and Droplets During High-Velocity Nasal Insufflation Therapy Using a Simple Surgical Mask: Implications for the High-Flow Nasal Cannula. Chest. 2020.

27. Leung CCH, Joynt GM, Gomersall CD, Wong WT, Lee A, Ling L, et al. Comparison of high-flow nasal cannula versus oxygen face mask for environmental bacterial contamination in critically ill pneumonia patients: a randomized controlled crossover trial. Journal of Hospital Infection. 2019;101(1):84–7.

28. El-Boghdadly K, Wong DJN, Owen R, Neuman MD, Pocock S, Carlisle JB, et al. Risks to healthcare workers following tracheal intubation of patients with COVID-19: a prospective international multicentre cohort study. Anaesthesia. 2020.

29. Chen WQ, Ling WH, Lu CY, Hao YT, Lin ZN, Ling L, et al. Which preventive measures might protect health care workers from SARS? BMC Public Health. 2009;9:81.

30. Liu W, Tang F, Fang L-Q, De Vlas SJ, Ma H-J, Zhou J-P, et al. Risk factors for SARS infection among hospital healthcare workers in Beijing: a case control study. Tropical Medicine & International Health. 2009;14(1):52–9.

31. Loeb M, McGeer A, Henry B, Ofner M, Rose D, Hlywka T, et al. SARS among critical care nurses, Toronto. Emerg Infect Dis. 2004;10(2):251–5.

32. Pei LY, Gao ZC, Yang Z, Wei DG, Wang SX, Ji JM, et al. Investigation of the influencing factors on severe acute respiratory syndrome among health care workers. Beijing da xue xue bao Yi xue ban = Journal of Peking University Health sciences. 2006;38(3):271–5.

33. Raboud J, Shigayeva A, McGeer A, Bontovics E, Chapman M, Gravel D, et al. Risk factors for SARS transmission from patients requiring intubation: a multicentre investigation in Toronto, Canada. PloS one. 2010;5(5):e10717–e.

34. Teleman MD, Boudville IC, Heng BH, Zhu D, Leo YS. Factors associated with transmission of severe acute respiratory syndrome among health-care workers in Singapore. Epidemiol Infect. 2004;132(5):797–803.

35. Hui DS, Hall SD, Chan MT, Chow BK, Tsou JY, Joynt GM, et al. Noninvasive positive-pressure ventilation: An experimental model to assess air and particle dispersion. Chest. 2006;130(3):730–40.

36. Hui DS, Chan MT, Chow B. Aerosol dispersion during various respiratory therapies: a risk assessment model of nosocomial infection to health care workers. Hong Kong Med J. 2014;20 Suppl 4:9–13.

37. Loh N-HW, Tan Y, Taculod J, Gorospe B, Teope AS, Somani J, et al. The impact of high-flow nasal cannula (HFNC) on coughing distance: implications on its use during the novel coronavirus disease outbreak. Can J Anaesth. 2020.

38. Fowler RA, Guest CB, Lapinsky SE, Sibbald WJ, Louie M, Tang P, et al. Transmission of Severe Acute Respiratory Syndrome during Intubation and Mechanical Ventilation. American journal of respiratory and critical care medicine. 2004;169(11):1198–202.

39. Ma HJ, Wang HW, Fang LQ, Jiang JF, Wei MT, Liu W, et al. [A case-control study on the risk factors of severe acute respiratory syndromes among health care workers]. Zhonghua Liu Xing Bing Xue Za Zhi. 2004;25(9):741–4.

40. Scales DC, Green K, Chan AK, Poutanen SM, Foster D, Nowak K, et al. Illness in intensive care staff after brief exposure to severe acute respiratory syndrome. Emerging infectious diseases. 2003;9(10):1205–10.

41. Wong T-w, Lee C-k, Tam W, Lau JT-f, Yu T-s, Lui S-f, et al. Cluster of SARS among medical students exposed to single patient, Hong Kong. Emerging infectious diseases. 2004;10(2):269–76.

42. Lamé G, Dixon-Woods M. Using clinical simulation to study how to improve quality and safety in healthcare. BMJ Simul Technol Enhanc Learning. 2020;6(2):87–94.

43. Fowler RA, Guest CB, Lapinsky SE, Sibbald WJ, Louie M, Tang P, et al. Transmission of severe acute respiratory syndrome during intubation and mechanical ventilation. Am J Respir Crit Care Med. 2004;169(11):1198–202.

44. Raboud J, Shigayeva A, McGeer A, Bontovics E, Chapman M, Gravel D, et al. Risk factors for SARS transmission from patients requiring intubation: a multicentre investigation in Toronto, Canada. PLoS One. 2010;5(5):e10717.

45. Balshem H, Helfand M, Schünemann HJ, Oxman AD, Kunz R, Brozek J, et al. GRADE guidelines: 3. Rating the quality of evidence. Journal of Clinical Epidemiology. 2011;64(4):401–6.

46. Ng K, Poon BH, Kiat Puar TH, Shan Quah JL, Loh WJ, Wong YJ, et al. COVID-19 and the Risk to Health Care Workers: A Case Report. Annals of internal medicine. 2020;172(11):766–7.

47. Heinzerling A, Stuckey PMJ, Scheuer T, Xu K, Perkins KM, Resseger H, et al. Transmission of COVID-19 to health care personnel during exposures to a hospitalized patient—Solano County, California, February 2020. 2020.

48. Feldman O, Meir M, Shavit D, Idelman R, Shavit I. Exposure to a surrogate measure of contamination from simulated patients by emergency department personnel wearing personal protective equipment. Jama. 2020;323(20):2091–3.

49. Ferreyro BL, Angriman F, Munshi L, Del Sorbo L, Ferguson ND, Rochwerg B, et al. Association of Noninvasive Oxygenation Strategies With All-Cause Mortality in Adults With Acute Hypoxemic Respiratory Failure: A Systematic Review and Meta-analysis. JAMA. 2020.

